# Phenotypically independent profiles relevant to mental health are genetically correlated

**DOI:** 10.1101/2020.03.30.20045591

**Authors:** Daniel Roelfs, Dag Alnæs, Oleksandr Frei, Dennis van der Meer, Olav B. Smeland, Ole A. Andreassen, Lars T. Westlye, Tobias Kaufmann

## Abstract

Genome-wide association studies (GWAS) and family-based studies have revealed partly overlapping genetic architectures between various psychiatric disorders. Given clinical overlap between disorders, our knowledge of the genetic architectures underlying specific symptom profiles and risk factors is limited. Here, we aimed to derive distinct profiles relevant to mental health in healthy individuals and to study how these genetically relate to each other and to common psychiatric disorders. Using independent component analysis, we decomposed self-report mental health questionnaires from 136,678 healthy individuals of the UK Biobank, excluding data from individuals with a diagnosed neurological or psychiatric disorder, into thirteen distinct profiles relevant to mental health, capturing different symptoms as well as social and risk factors underlying reduced mental health. Utilizing genotypes from 117,611 of those individuals with White English ancestry, we performed GWAS for each mental health profile and assessed genetic correlations between these profiles, and between the profiles and common psychiatric disorders and cognitive traits. We found that mental health profiles were genetically correlated with a wide range of psychiatric disorders and cognitive traits, with strongest effects typically observed between a given mental health profile and a disorder for which the profile is common (e.g. depression symptoms and major depressive disorder, psychosis and schizophrenia). Strikingly, although the profiles were phenotypically uncorrelated, many of them were genetically correlated with each other. This study provides evidence that statistically independent mental health profiles partly share genetic underpinnings and show genetic overlaps with psychiatric disorders, suggesting that shared genetics across psychiatric disorders cannot be exclusively attributed to the known overlapping symptomatology between the disorders.

## Introduction

Psychiatric disorders are highly polygenic, exhibiting a multitude of significantly associated genetic variants with small effect sizes. Recent large-scale genome-wide association studies (GWAS) have identified a large number of single-nucleotide polymorphisms (SNP) associated with psychiatric disorders such as schizophrenia (SCZ)^1^, bipolar disorder (BD)^2^, major depression (MD)^3^, attention deficit hyperactivity disorder (ADHD)^4^, autism spectrum disorders (ASD)^5^, post-traumatic stress disorder (PTSD)^6^, and anxiety (ANX)^7^. In addition to substantial polygenicity, previous findings have documented genetic overlap between disorders^8–11^, even in the absence of genetic correlations expressed as additive genetic effects for two traits, as recently demonstrated for schizophrenia and educational attainment^12,13^. Adding to the complexity, psychiatric disorders also overlap with multiple complex traits, such as BMI^14^, cardio-metabolic diseases^15^ and a number of psychosocial and other risk factors for reduced mental health^16–18^. The latter are particularly challenging in the context of genetics, since genetic overlap may not necessarily point to causative effects but rather point at common underlying factors^19,20^. Taken together, the landscape of current psychiatric genetics suggests highly complex patterns of associations and unclear specificity for many common psychiatric disorders.

While GWAS studies have allowed to disentangle parts of the genetic architecture of psychiatric disorders, these methods alone are not sufficient to answer some of the challenges posed in psychiatric genetics. One of those challenges is the lack of clinical demarcation between psychiatric disorders. For example, patients with the same diagnosis may not necessarily exhibit common symptoms^21^ and patients with different diagnoses may show highly overlapping clinical phenotypes^22^. The notion that mental disorders like schizophrenia and bipolar disorders reflect biologically heterogeneous categories is also supported by neuroimaging studies^23,24^. Nonetheless, a majority of large-scale genetic studies use a classical case-control design based on a categorical operationalization of disease without stratifying other measures such as symptoms, functioning or symptom severity. Likewise, control groups are rarely screened for subthreshold symptoms. For example in the case of psychosis, approximately 6% of the general population are reported to have a psychotic experience in their lifetime, and only a minority of that group will develop a diagnosed psychiatric illness such as schizophrenia or bipolar disorder^25^. Finally, the likelihood of inducing selection bias when drawing cases and controls from different populations are high and may impose confounds in case-control designs^26^. Thus, whereas studies using the classical case-control design have been instrumental and produced a strong body of discoveries in psychiatric genetics, these designs have limitations that may prevent us from discovering signal more closely related to clinical characteristics of the disorder. In addition, case-control designs require immense effort and resources given that the high polygenicity of common psychiatric disorders requires vast sample sizes to detect effects^27^.

Recent large-scale population level efforts such as the UK Biobank^28^ now provide alternatives for the study of psychiatric disorders. The mental health data available in UK Biobank includes data from more than 150,000 individuals and covers questions on current and previous symptoms in different psychiatric domains^29^. For example, a recent study revealed genetic associations with psychotic experiences in the UK Biobank and reported genetic correlations between psychotic experience and common psychiatric disorders^30^. While this study formed two groups of subjects (with and without psychotic experience) others have suggested continuous measures of psychopathology obtained from questionnaire data, such as the p-factor^31^. While bundling variance of psychopathology in a single common factor can be a useful proxy of mental health vulnerability, the specificity of the p-factor to disorder-specific mechanisms is limited^31^. Independent component analysis^32,33^ provides a complementary approach to decompose the variance from mental health questionnaires into independent latent variables. For example, using independent component analysis on mental health questionnaires of children and adolescents, Alnæs and colleagues have identified a set of independent components reflecting symptoms of attention deficit, psychosis, depression, anxiety, and more^34^. Independent components obtained from mental health questionnaires may each capture either global (e.g. joint symptoms of depression, stress and anxiety) or specific aspects (e.g. pure psychosis symptoms) of mental health in a data-driven fashion, thereby yielding multiple distinct profiles relevant to mental health symptoms beyond a common p-factor.

Here, in order to disentangle the genetic architecture underlying psychiatric symptoms and traits as well as psychosocial and other risk factor for reduced mental health we investigated structures of psychopathology and corresponding genetics using independent component analysis in the UK Biobank mental health data. This allowed us to study the genetic relationships between statistically independent profiles relevant to mental health, and between these profiles and psychiatric disorders as well as cognitive traits. We focused our analysis on data from individuals who had no previous diagnosis with a neurological or psychiatric disorder, yielding novel insights into variation in mental health in a healthy (undiagnosed) population. Given that preclinical symptoms in healthy individuals may share biological mechanisms with symptoms in diagnosed individuals, we hypothesized that the genetic architecture of specific variations in mental health in *healthy* (undiagnosed) individuals overlaps with specific major psychiatric disorders. However, we did not have an a-priori hypothesis for the degree of specificity. The known pleiotropy between major psychiatric disorders (reproduced in Suppl. Fig. 1) might reflect similar symptoms or risk factors occurring in different disorders or similar mechanisms underlying different symptoms or risks. We therefore investigated if statistically independent mental health profiles are also genetically independent or if they share a common genetic architecture, which may yield insights into the sources of pleiotropy in psychiatric genetics.

## Methods and Materials

### Sample and exclusion criteria

We accessed data from the UK Biobank^28^ with permission no. 27412, and included data from individuals who had participated in an online follow-up questionnaire on mental health (UK Biobank category 136). All participants provided signed informed consent before inclusion in the study. UK Biobank was approved by the National Health Service National Research Ethics Service (ref. 11/NW/0382). Participants with a diagnosed psychiatric or neurological disorder (F or G ICD10 diagnosis) were excluded from the analysis except for those with a nerve, nerve root and plexus disorders (categories G50 to G59). In addition, we excluded participants with more than 10% missing answers in the mental health questionnaires. This resulted in mental health data from 136,678 individuals, which was used in an independent component analysis. For the genetic analysis, we selected data from all White English individuals with available genotypes, yielding a set of 117,611 participants aged 47-80 years (mean: 64, SD: 7.66, age at mental health assessment) and comprised 56.2% females.

### Processing of mental health data

Fig. 1A depicts the analysis workflow. The UK Biobank database contains about 140 questions on mental health and risk factors related to reduced mental health. The questions on the mental health risk factors are retrospective (e.g. “have you ever …”, “at any point in your lifetime …” etc.). This warrants some caution in the interpretation, since retrospective analysis yields lower power than a prospective design. We selected only primary questions that were answered by all participants, excluding follow-up questions. Of the 60 resulting questions that were available, we removed questions that asked specifically about symptoms occurring in the past two weeks to remove potential short-term temporal effects. Furthermore, we excluded questions where more than 10% of the responses were missing (1 question excluded). In the resulting set of 43 questions (Suppl. Table 1), we imputed missing data using k-nearest neighbor imputation with k=3 with the *bnstruct* package^35^ in R^36^ and z-standardized the data (Suppl. Fig. 2).

**Fig. 1.**
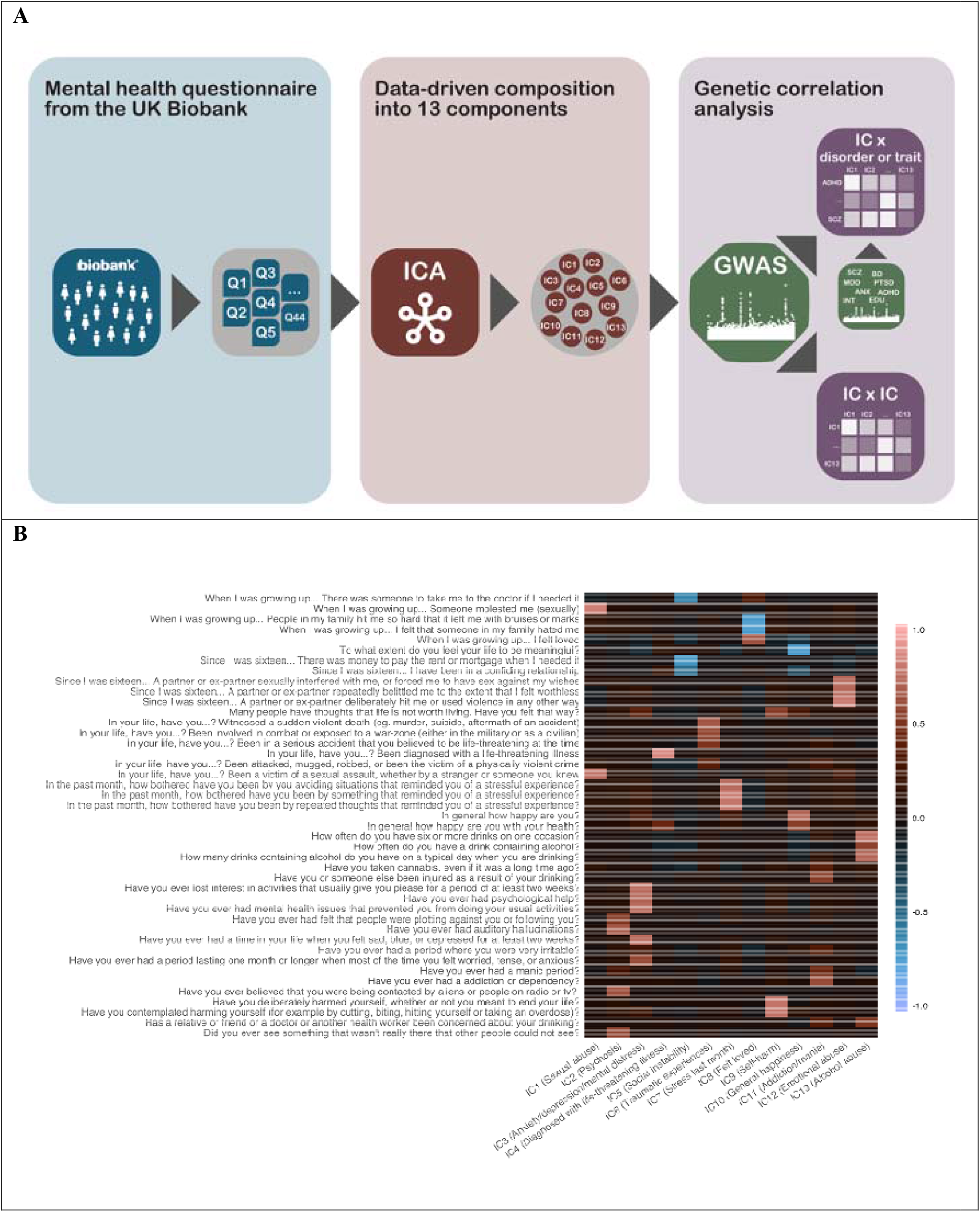
Workflow and variable weight matrix of the resulting decomposition. **A** Outline of the analysis workflow. **B** Weight matrix reflecting how each mental health question loaded on each IC. Brighter colors indicate higher loading, darker colors indicate lower loading. All 43 questions were captured by at least one of the 13 independent components. To facilitate interpretation, loadings of IC1, IC2, IC5, IC9, IC10, IC11, and IC12 were inverted so that all components showed the same direction of effect (higher component score indicating increased scoring on the items).

The resulting data covering 43 questions from 136,678 individuals was decomposed using independent component analysis (ICA). ICA is a statistical clustering method that decomposes multiple related variables into statistically independent components. The resulting components show a high degree of within-cluster correlation, but no correlation between the clusters. Of note, the number of components needs to be pre-specified, and this selection of appropriate model order is to a certain degree a subjective task where depending on the stringency of the criteria defined for model order selection it is possible to obtain several different solutions that meet the requirements for an appropriate threshold. Here, we used icasso^37^ in MATLAB in combination with visual inspection of the loadings of the questions on the components. The PCA identified 13 components with an eigenvalue larger than 1, and stability (Iq) was effectively 1. A model order lower than 13 would group together questions into components which we preferred to keep separate. A model order larger than 13 was not reasonable as it would yield components that largely reflect single items. We therefore concluded that a model order of 13 independent components yields the best clustering solution where the resulting components are stable and highly interpretable. The individual scores for each of the 43 questions were subsequently residualized for age (both linear and quadratic term), sex, and the first 20 genetic principal components. Next, we decomposed the items into 13 independent components using the fastICA algorithm as implemented in R^38^. Fig. 1B depicts how each of the 43 items loaded on the components, indicating independent components (ICs) that captured questions on sexual abuse (IC1), psychosis (IC2), anxiety, depression and mental distress (IC3), a diagnosis with a life-threatening illness (IC4), social instability (IC5), traumatic experiences (IC6), stress in the past month (IC7), experiences of feeling loved (IC8), thoughts around self-harm behavior (IC9), general happiness (IC10), addiction behavior and manic experiences (IC11), experiences of emotional abuse (IC12), and alcohol abuse (IC13). Of note, we here introduced this labeling of the ICs only to improve legibility of the results yet caution is warranted as the label is not necessarily encompassing all facets of a given component. The labels only highlight some of the core domains of questions weighing strongly on a given components, yet all interpretations need to be made in the light of the ICA framework (Fig. 1B)

The distribution of IC2 indicated very few non-zero scores (Suppl. Fig. 3). This component loaded mostly on psychosis questions (Fig. 1B), indicating that only few of the included healthy individuals had symptoms in this domain. We therefore conducted an additional supplemental analysis in which we dichotomized IC2 such that loadings lower than 1 were labeled as “no/few symptoms”, and loadings equal to or higher than 1 were labeled as “with symptoms”.

### Processing of genetic data

From the UK Biobank v3 imputed genetic data, we removed SNPs with an imputation quality score below 0.5, with a minor allele frequency below 0.001, missing in more than 5% of individuals, and that failed the Hardy-Weinberg equilibrium test at p < 1e-9. We removed also individuals with more than 10% missing data. We performed a genome-wide association analysis (GWAS) on each of the 13 independent components in PLINK 2^39,40^. Using a publicly available conversion toolbox for GWAS summary statistics (github.com/precimed/python_convert), we removed the MHC region and calculated a z-score for every SNP (8,165,726 SNPs after QC). We utilized linkage-disequilibrium score regression^10,41^ to estimate genetic correlations between each of the independent components, and between the components and publicly available GWAS summary statistics for SCZ^1^, BD^2^, MD^42^, ADHD^4^, ASD^5^, PTSD^6^, ANX^7^, as well as intelligence^43^, and educational attainment^44^ (Suppl. Table 2). For all aforementioned GWASs, we used those versions that did not have UK Biobank participants included. From the MD GWAS, we removed participants from the 23andMe dataset as well, leaving only cases with a diagnosed major depressive disorder (MDD). Prior to estimating genetic correlations, we set a threshold that only ICs with a heritability 1.96 times larger than its standard error should be included in the analysis and only those where visual quality control of corresponding Q-Q plots indicated genetic signal. These quality control steps were implemented to ensure that we did not make inferences on data that did not provide sufficient variance explained by genetics. Partitioned heritability^45^ was estimated using the LDSC toolbox^41^ and Q-Q plots were generated using custom scripts in R. Finally, we processed the GWAS summary statistics of each independent component through the Functional Mapping and Annotation toolbox (FUMA) to map lead SNPs onto genes^46^. FUMA parameters were kept as default, and we used the FUMA default European ancestry reference panel.

### Code and data availability

Code and GWAS summary statistics will be made publicly available via GitHub (github.com/norment) upon acceptance of the manuscript. Furthermore, the derived independent components (individual level data) will be made available to the UK Biobank upon acceptance (derived variable return) to allow its use in future UK Biobank studies.

## Results

Fig. 2 depicts SNP-based heritability (h2) for the 13 ICs (Suppl. Table 3 for additional statistics). Heritability was generally low, yet all components yielded a heritability that was higher than 1.96 times the standard error. IC13, capturing questions on alcohol abuse had the highest heritability (h2 = 0.0763, SE = 0.0055), closely followed by IC3, capturing anxiety, depression, and mental distress (h2 = 0.0744, SE = 0.0052). The lowest heritability among the components was for IC2, reflecting psychosis questions (h2 = 0.0089, SE = 0.0043), likely owing to the low number of individuals with psychosis symptoms (Suppl. Fig. 3). We therefore performed a supplemental analysis to investigate if dichotomization of this IC would benefit the analysis (Suppl. Fig. 4). In brief, as dichotomization only slightly improved heritability estimates, we kept IC2 as a continuous component for the main analysis to stay consistent with the other components, yet we provide results with the dichotomized component in Suppl. Fig. 4. In addition to passing the heritability criterion of 1.96 times the standard error, the Q-Q plots of all ICs passed visual quality control (Suppl. Fig. 5) warranting inclusion of all components into subsequent genetic correlation analyses.

**Fig. 2.**
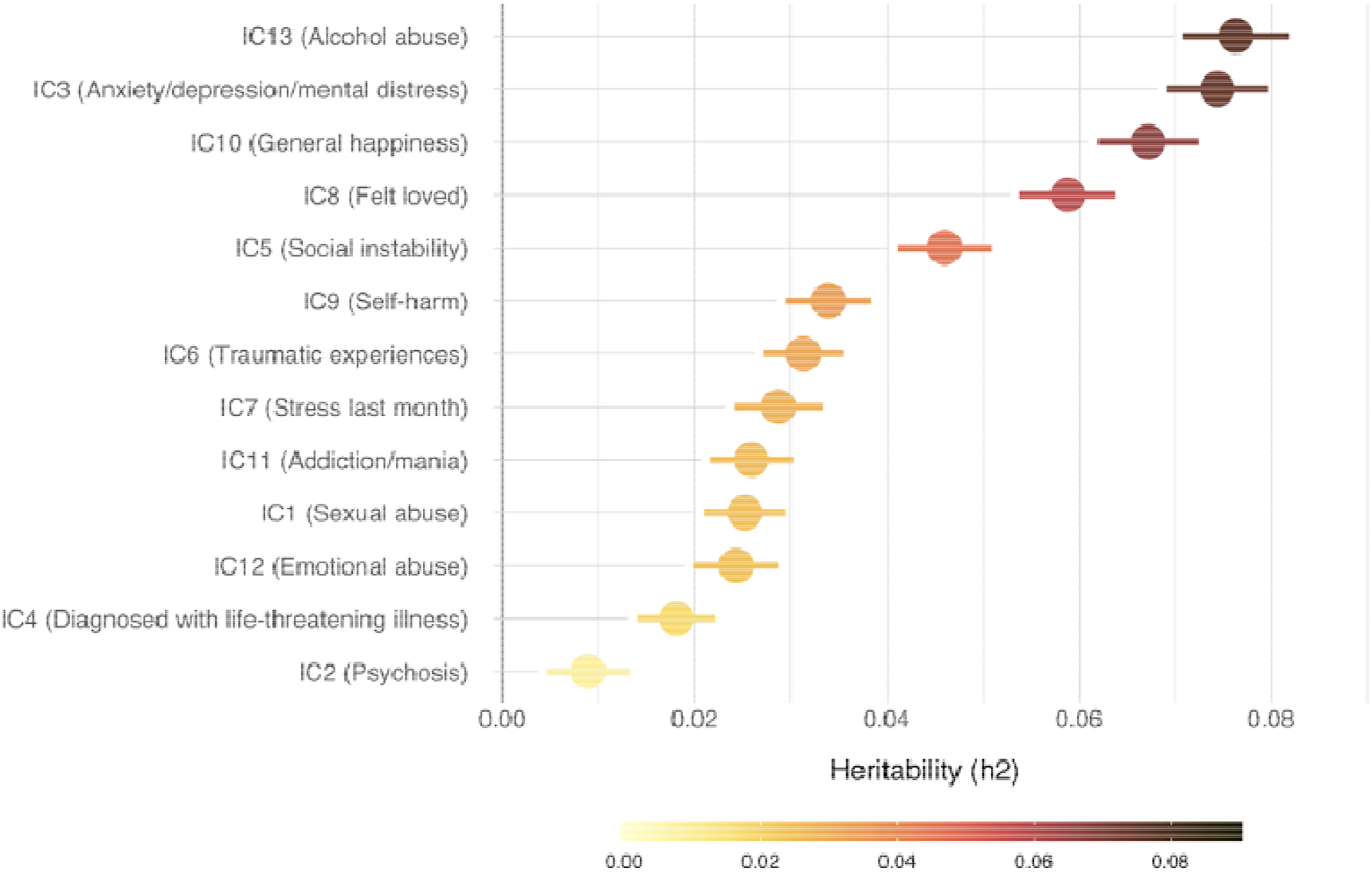
Heritability estimates of the independent components. SNP-based heritability for each IC sorted by decreasing heritability (h2). Heritability calculated using the LDSC toolbox^41^. Error bars reflect standard errors.

Except for IC4, all ICs showed genome wide significant SNPs at a threshold of 5e-8 (Suppl. Fig. 6). Using FUMA, we discovered 7 independent loci for IC13, 2 for IC2, IC7, and IC8, 1 locus for IC1, IC3, IC5, IC10, IC11 and IC12, and IC4, IC6, and IC9 had no significant genetic risk loci. Suppl. Table 4 provides a list of mapped genes for all ICs, illustrating that IC13 had the most mapped genes among all ICs (74 mapped genes).

We assessed genetic correlations between each of the 13 ICs and a set of psychiatric disorders as well as cognitive traits. Out of 117 comparisons, 70 were significant after FDR correction (α = 0.05), which amounts to 59%. Fig. 3 depicts all genetic correlations with ICs, sorted separately for each disorder or cognitive trait (sorted by absolute genetic correlation). Suppl. Fig. 7 shows the same genetic correlations separated by IC. We found that in most cases the strongest genetic correlation was with the IC most closely related to that disorder or trait. For example, anxiety most strongly correlated with IC3, which reflects anxiety, depression, and mental distress (genetic correlation rg = 0.70, pFDR < .00027). SCZ was most highly correlated with IC2, which represents psychosis questions (rg = 0.54, pFDR = .001). The highest genetic correlation of BD was with IC11, which represents addiction and mania (rg = 0.5, pFDR = 6.5e-12). For PTSD, the component reflecting traumatic experience (IC6) only ranked sixth among the sorted associations, yet the two ICs showing strongest association with PTSD reflected anxiety, depression, and mental distress (IC3; rg = 0.53, pFDR = .0017) and diagnosed with life-threatening illness (IC4; rg = 0.51, pFDR = .080), both of which are closely related to PTSD. ASD correlated strongest with IC2 (reflecting psychosis; rg = 0.40, pFDR = .031) and ADHD correlated strongest with IC8 (Felt loved; rg = −0.51, pFDR = 4.7e-21). Educational attainment and intelligence were both strongest negatively correlated with the IC reflecting social instability (IC5, rg = −0.74 and rg = −0.76, respectively; both pFDR < 2.5e-74). In general, the strongest associations among all ICs, either positive or negative were with MDD while the weakest associations were with educational attainment.

**Fig. 3.**
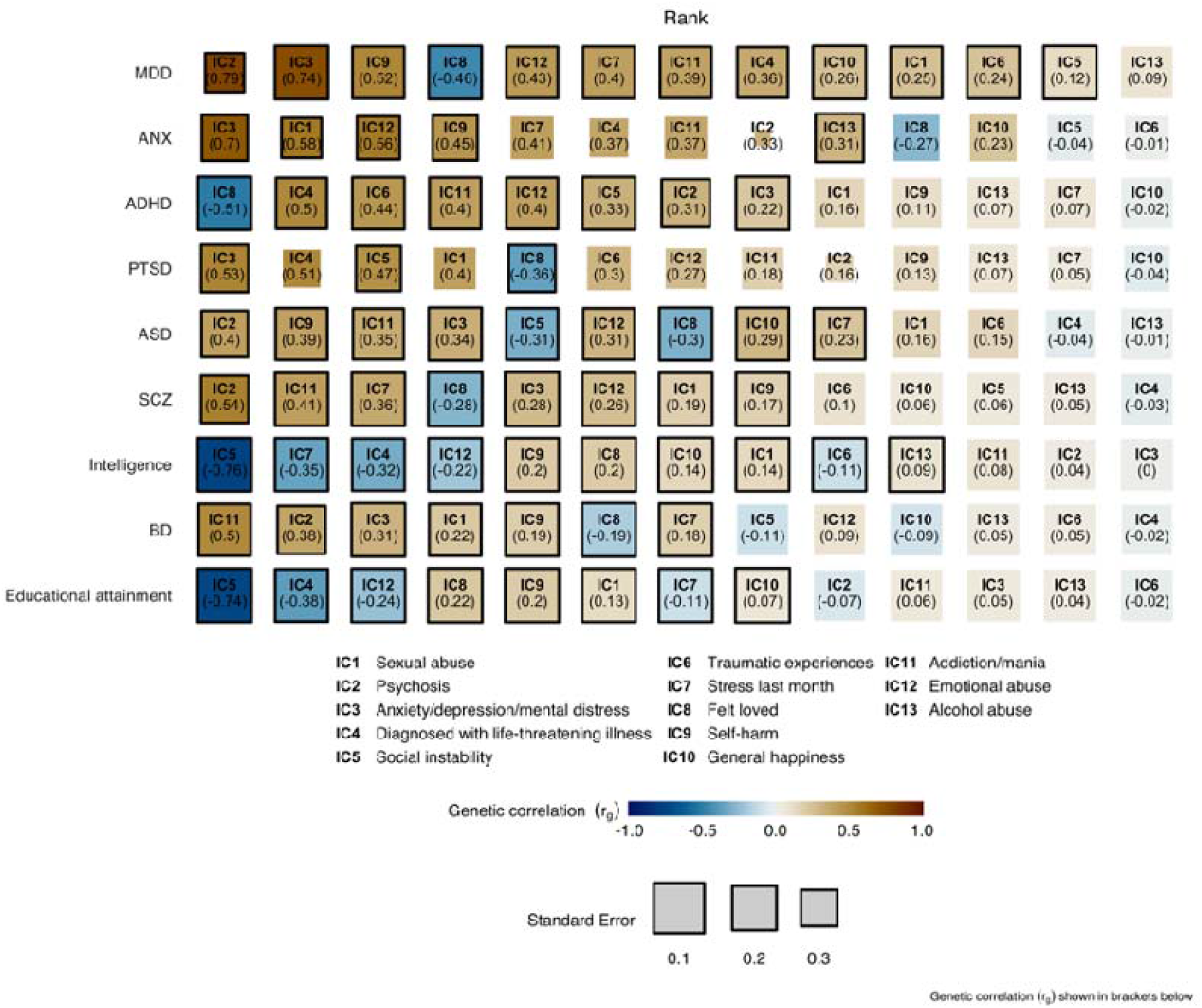
Genetic correlation between the independent components and disorders and cognitive traits. For each disorder, the associations with ICs are sorted by decreasing absolute genetic correlation such that the most leftward box reflects the strongest association between a given disorder and the 13 ICs. Numbers in brackets under each IC label denote the genetic correlation (rg). Size of the boxes reflect the standard error. Significant correlations (p < FDR) are indicated with a black border.

Next, we assessed the genetic correlations between the ICs. Independent components are statistically independent by design, and thus on the phenotype level the ICs were not correlated with each other (Fig. 4, lower half; correlations essentially zero). However, almost half of the IC pairs were nonetheless significantly genetically correlated with each other (45%, p < FDR). IC3 (anxiety, depression, mental illness) was genetically correlated with 10 other ICs. IC9 (self-harm) was correlated with 9 other ICs and IC6 (traumatic experiences) and IC8 (felt loved) were each genetically correlated with eight other ICs. IC11 (addiction/mania) and IC12 (emotional abuse) were each genetically correlated with seven other ICs. IC1 (sexual abuse) and IC5 (social instability) were both genetically correlated with six other ICs. IC2 (psychosis) was correlated with 5 other ICs. IC4 (diagnosed with life-threatening illness) and IC13 (alcohol abuse) were both genetically correlated with 4 other ICs. And IC7 (stress last month) and IC10 (general happiness) were both genetically correlated with 3 other ICs. No IC had no significant genetic correlations with other ICs. The analysis therefore revealed a large amount of genetic correlations despite statistical (phenotypic) independence of the symptom profiles.

**Fig. 4.**
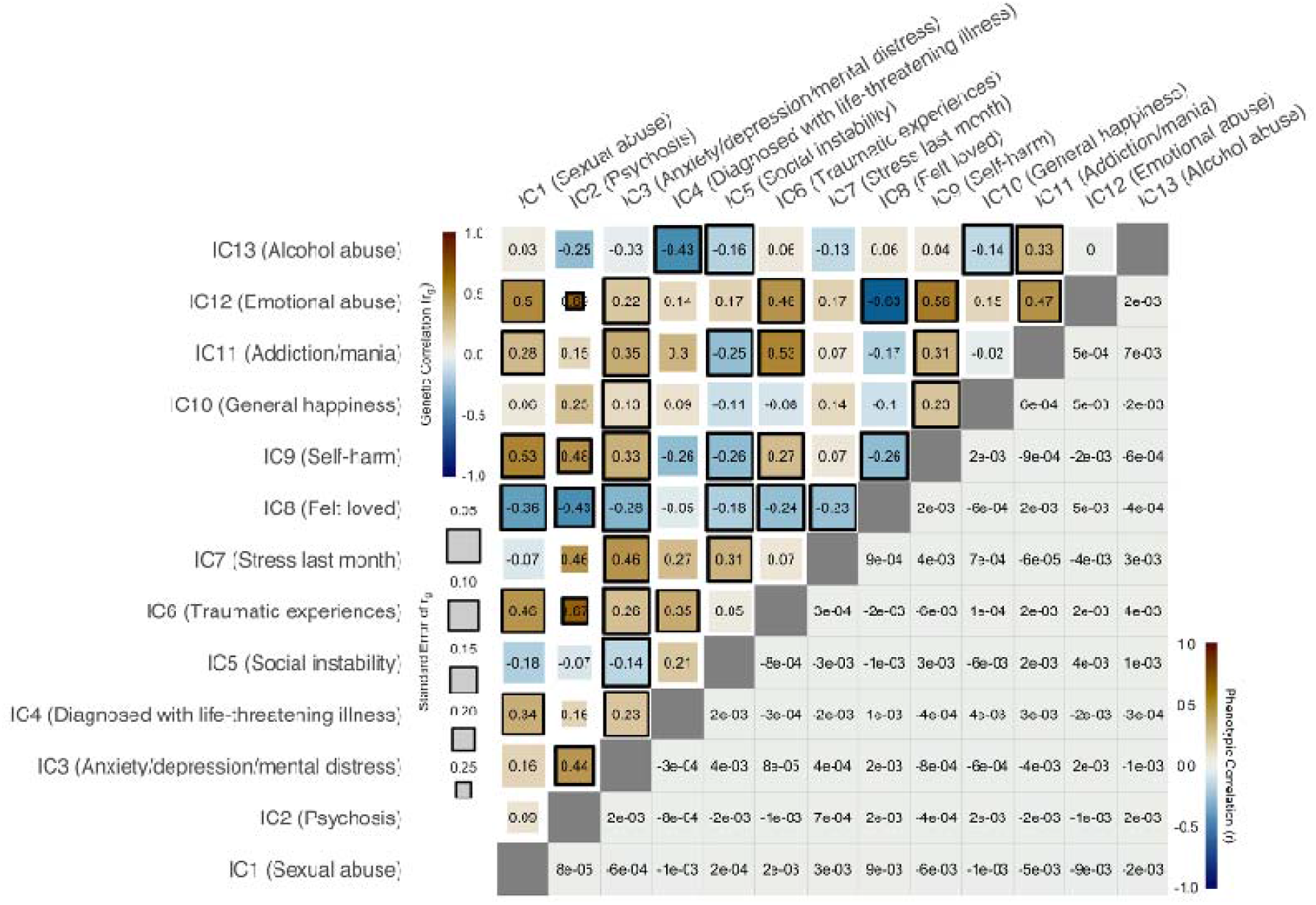
Phenotypic and genetic correlation between the ICs. The lower half of the IC by IC matrix depicts phenotypic correlations, reflecting the Pearson correlation of subject level component scores between independent components. As expected by ICA design, correlations were close to zero. The upper half of the matrix depicts the genetic correlations (rg), indicating significant genetic correlations in 40 of 78 tests. Size of the boxes indicate standard error and significant correlations (p < FDR) are indicated with a black border.

## Discussion

In the present study, we decomposed mental health questionnaire data from more than 130,000 individuals into phenotypically distinct profiles relevant to mental health (independent components) that reflected compositions of symptoms, psychosocial and other risk factors for reduced mental health. We found that variations in these profiles in healthy individuals (without a neurological or psychiatric diagnosis) were genetically correlated with psychiatric disorders and cognitive traits. Strongest correlations were observed between components and disorders with known symptoms in a similar domain (e.g. psychosis symptoms with schizophrenia), but the large amount of significant correlations between disorders and mental health profiles suggested limited specificity. Indeed, we found a large proportion of significant genetic correlations between the phenotypically uncorrelated profiles, suggesting overlapping genetic architectures underlying distinct symptoms and risk factors. A number of the questions included in the analyses revolved around risk factors for mental health, such as a history of childhood abuse, sexual abuse, and an unstable home situation. Caution is warranted in the interpretation of these effects. The genetic correlation with the independent components capturing these items do not suggest that there is a genetic component to high-risk environments but rather are likely to capture second order effects. In other words, a GWAS on such these risk factors is more likely to reflect other factors underlying these risk factors than the risk factor directly.

The implications of our findings are twofold. First, our results support pleiotropy in psychiatric disorders beyond overlapping symptoms (e.g. BD and MDD both involving depressive episodes), suggesting that even distinct psychiatric symptoms are genetically overlapping. Second, our findings support that normal variability in mental health within healthy individuals may inform the study of the biology of psychiatric disorders.

While pleiotropy between major psychiatric disorders has been widely established^9–11^ (reproduced in Suppl. Fig. 1), the sources underlying pleiotropy remain largely unknown. Specifically, disorders oftentimes overlap in symptomatology and therefore the degree to which the observed genetic correlations between disorders reflect phenotypic overlap between disorders remains to be investigated. Our approach of decomposing mental health data into distinct profiles allowed us to study genetic correlations in a sample with known phenotypic correlations and to assess how these profiles correlate with the genetics of different diagnoses. We observed that most disorders correlated strongest with the independent components capturing a related phenotype. For example, the strongest association with IC3, which reflects variance in anxiety, depression, and mental distress, was with ANX, the strongest association with IC2 (psychosis) was with SCZ. Therefore, the ranking of association strengths suggested a certain degree of specificity. However, that degree was strongly limited as most of the disorders and components were significantly genetically correlated. For example, MDD showed significant correlations between all but one component, ASD correlated with all but four components, and ANX and ADHD were correlated with all but 5 components, though correlation strengths were overall lower than with MDD, possibly due to lower sample size. There were also significant associations between components and cognitive traits although overall weaker associations compared to those with disorders. About half of the genetic correlations with intelligence and educational attainment pointed in the opposite direction, considerably more than for the psychiatric disorders, reflecting higher cognitive ability with fewer psychiatric symptoms. Importantly, when looking at the correlations between mental health profiles, we found that almost half of the genetic correlation matrix between ICs yielded significant genetic correlations despite a lack of phenotypic correlations (independence of the components). This suggests that some of the same genes are involved in the genetics of distinct profiles relevant to mental health and may indirectly support pleiotropy independent of phenotypic overlap in psychiatric disorders. Whereas more research is needed before conclusions on the sources underlying the observed pleiotropy can be drawn, one possible explanation for the significant correlations in the ICs could be that, since all independent components each capture a facet of mental health, there may be a number of SNPs that are involved across mental health symptoms. These SNPs may be involved in overall mental health, from psychological well-being to psychosis symptoms. Our analysis of significant SNPs in FUMA did not identify overlapping SNPs between ICs, however, this may be attributed to the relatively low number of significant loci discovered in the ICs. Advanced statistical tools and further increasing sample sizes may help pinpoint specific genes involved with different symptoms. Furthermore, it is also plausible that environmental effects may factor into the explanation of the significant genetic correlations despite phenotypic independence if the environmental factors differ markedly between the ICs.

### Limitations

Notable strengths of the present study include the use of data-driven decomposition of mental health data in a large sample of healthy individuals and its application to study pleiotropy in psychiatric genetics. Its main limitations include the low heritability of the resulting independent components, and the limited number of individuals with psychosis symptoms yielding suboptimal distribution in IC2 (Suppl. Fig. 3). First, it is important to note that all ICs passed quality control. Heritability of all ICs exceeded our pre-defined heritability threshold of 1.96 times its standard error, and all Q-Q plots indicated genetic signal (Suppl. Fig. 5). Furthermore, low heritability can still produce good genetic signal as a result from a low number of genetic variants involved but where each has large effects^13^. For example, while IC2 had the lowest heritability among the ICs, it showed one of the strongest genetic signals and together with IC7 and IC8 it ranked second in terms of the number of loci discovered in FUMA, following IC13 (alcohol abuse) that showed the highest heritability, strongest genetic signal on the Q-Q plot and the largest number of significant loci and mapped genes. Second, although sample size and symptom distributions factored into the results, these are mostly reflected in the standard error of genetic associations, not in a lack of effect. For example, ANX^7^ (n = 21,761) and PTSD^6^ (n = 9,537) GWASs have relatively little power as reflected in the larger standard errors in genetic correlations with these disorders, but nonetheless the strongest associations with these disorders were with components that match symptoms of the disorders (both correlated strongest with IC3, reflecting anxiety/depression/mental distress). Likewise, the suboptimal symptom distributions in IC2 and corresponding low heritability is reflected in large standard errors of the resulting genetic correlations but nonetheless IC2, reflecting psychosis, was most strongly associated with SCZ. Supplemental analysis with dichotomized IC2 also confirmed that the distribution alone is unlikely to explain the observed associations (Suppl. Fig. 4).

Furthermore, it is important to note that while we excluded individuals with psychiatric disorders based on ICD codes we may still include individuals with (sub-threshold) psychiatric disorders that have not been diagnosed. For example, most patients with depressive symptoms in the UK will be treated by first-line care, which may not be registered in the UK Biobank^47^. However, distributions of z-scores on the individual questions appear quite similar between individuals without and individuals with a diagnosis (Suppl. Fig. 2). While we cannot rule out subtle contributions on IC decomposition by the potential remaining inclusion of a subset of patients in primary care, these results support that should such a confound be present, it is unlikely to have introduced enough structural variance to diminish our main findings.

### Conclusion

In the present study, we revealed genetic overlap between statistically independent profiles relevant to mental health capturing compositions of symptoms, psychosocial and other risk factors for reduced mental health and provide evidence that variations in mental health in healthy individuals relate genetically to psychiatric disorders and cognitive traits. These findings support that pleiotropy between psychiatric disorders cannot simply be explained by overlapping symptoms or risks but may rather point to similar biological underpinnings of distinct symptoms or risks. Our results underscore the potential of data-driven approaches to the study of mental health, and suggests that supplementing the classic case-control design with a dimensional approach may improve our understanding of the genetic underpinnings of complex disorders of the mind.

## Acknowledgements

The authors were funded by the Research Council of Norway (#276082 LifespanHealth, #223273 NORMENT, #283798 ERA-NET Neuron SYNSCHIZ, #249795), the South-East Norway Regional Health Authority (2019101, 2019107, and 2020086), and the European Research Council under the European Union’s Horizon2020 Research and Innovation program (ERC Starting Grant #802998), as well as the Horizon2020 Research and Innovation Action Grant CoMorMent (#847776). This research has been conducted using the UK Biobank Resource (access code 27412, https://www.ukbiobank.ac.uk/). This work was performed on the TSD (Tjenester for Sensitive Data) facilities, owned by the University of Oslo, operated and developed by the TSD service group at the University of Oslo, IT-Department (USIT). Computations were also performed on resources provided by UNINETT Sigma2 - the National Infrastructure for High Performance Computing and Data Storage in Norway.

## Author contributions

D.R. and T.K. conceived the study; D.R. analyzed the data with contributions from T.K.; All authors contributed with conceptual input on methods and/or interpretation of results; D.R. and T.K. wrote the first draft of the paper and all authors contributed to the final manuscript.

