## Supplemental Material for "Phenotypically independent profiles relevant to mental health are genetically correlated"

**Supplementary Tables**

| **no** | **question** | **coding** |
| --- | --- | --- |
| 1 | In your life, did you seek or receive help from a professional (medical doctor, psychologist, social worker, counsellor, nurse, clergy, or other helping professional) for mental distress, psychological problems or unusual experiences? | 502 |
| 2 | In your life, have you suffered from a period of mental distress that prevented you from doing your usual activities? | 502 |
| 3 | Have you ever had a time in your life lasting two weeks or more when you lost interest in most things like hobbies, work, or activities that usually give you pleasure? | 503 |
| 4 | Have you ever had a time in your life when you felt sad, blue, or depressed for two weeks or more in a row? | 503 |
| 5 | Have you ever had a period of time when you were feeling so good, high, excited, or hyper that other people thought you were not your normal self or you were so hyper that you got into trouble? | 502 |
| 6 | Have you ever had a period of time when you were so irritable that you found yourself shouting at people or starting fights or arguments? | 502 |
| 7 | Have you ever had a period lasting one month or longer when most of the time you felt worried, tense, or anxious? | 502 |
| 8 | Have you been addicted to or dependent on one or more things, including substances (not cigarettes/coffee) or behaviours (such as gambling)? | 502 |
| 9 | In the next two questions, a drink is defined as one unit of alcohol. How many drinks containing alcohol do you have on a typical day when you are drinking? | 522 |
| 10 | Has a relative or friend or a doctor or another health worker been concerned about your drinking or suggested you cut down? | 524 |
| 11 | Have you or someone else been injured as a result of your drinking? | 524 |
| 12 | How often do you have a drink containing alcohol? | 521 |
| 13 | In the next two questions, a drink is defined as one unit of alcohol. How often do you have six or more drinks on one occasion? | 523 |
| 14 | Have you taken CANNABIS (marijuana, grass, hash, ganja, blow, draw, skunk, weed, spliff, dope), even if it was a long time ago? | 526 |
| 15 | Did you ever hear things that other people said did not exist, like strange voices coming from inside your head talking to you or about you, or voices coming out of the air when there was no one around? | 502 |
| 16 | Did you ever believe that there was an unjust plot going on to harm you or to have people follow you, and which your family and friends did not believe existed? | 502 |
| 17 | Did you ever see something that wasn't really there that other people could not see? | 502 |
| 18 | Did you ever believe that a strange force was trying to communicate directly with you by sending special signs or signals that you could understand but that no one else could understand (for example through the radio or television)? | 502 |
| 19 | When I was growing up... I felt that someone in my family hated me | 532 |
| 20 | When I was growing up... People in my family hit me so hard that it left me with bruises or marks | 532 |
| 21 | When I was growing up... I felt loved | 532 |
| 22 | When I was growing up... Someone molested me (sexually) | 532 |
| 23 | When I was growing up... There was someone to take me to the doctor if I needed it | 532 |
| 24 | Next is a list of problems and complaints that people sometimes have in response to such extremely stressful experiences. Please indicate how much you have been bothered by that problem in the past month: Avoiding activities or situations because they reminded you of a stressful experience? | 534 |
| 25 | Next is a list of problems and complaints that people sometimes have in response to such extremely stressful experiences. Please indicate how much you have been bothered by that problem in the past month: Repeated, disturbing memories, thoughts, or images of a stressful experience? | 534 |
| 26 | Next is a list of problems and complaints that people sometimes have in response to such extremely stressful experiences. Please indicate how much you have been bothered by that problem in the past month: Feeling very upset when something reminded you of a stressful experience? | 534 |
| 27 | Since I was sixteen... A partner or ex-partner repeatedly belittled me to the extent that I felt worthless | 532 |
| 28 | Since I was sixteen... I have been in a confiding relationship | 532 |
| 29 | Since I was sixteen... A partner or ex-partner deliberately hit me or used violence in any other way | 532 |
| 30 | Since I was sixteen... A partner or ex-partner sexually interfered with me, or forced me to have sex against my wishes | 532 |
| 31 | Since I was sixteen... There was money to pay the rent or mortgage when I needed it | 532 |
| 32 | In your life, have you...? Been in a serious accident that you believed to be life-threatening at the time | 533 |
| 33 | In your life, have you...? Been involved in combat or exposed to a war-zone (either in the military or as a civilian) | 533 |
| 34 | In your life, have you...? Been diagnosed with a life-threatening illness | 533 |
| 35 | In your life, have you...? Been attacked, mugged, robbed, or been the victim of a physically violent crime | 533 |
| 36 | In your life, have you...? Witnessed a sudden violent death (eg. murder, suicide, aftermath of an accident) | 533 |
| 37 | In your life, have you...? Been a victim of a sexual assault, whether by a stranger or someone you knew | 533 |
| 38 | Many people have thoughts that life is not worth living. Have you felt that way? | 535 |
| 39 | Have you deliberately harmed yourself, whether or not you meant to end your life? | 503 |
| 40 | Have you contemplated harming yourself (for example by cutting, biting, hitting yourself or taking an overdose)? | 535 |
| 41 | In general how happy are you? | 537 |
| 42 | In general how happy are you with your HEALTH? | 537 |
| 43 | To what extent do you feel your life to be meaningful? | 538 |
| **Suppl. Table 1.** **Full list of questions** This table lists the questions included in this study as presented to the participant. Data was available from 157,352 individuals, but through exclusion criteria 136.678 individuals remained. There are in total 43 questions. The third column indicates the coding. Possible answers for each coding are as follows:  502 – Prefer not to answer/Do not know/No/Yes  503 – Prefer not to answer/No/Yes  521 – Prefer not to answer/Never/Monthly or less/2 to 4 times a month/2 to 3 times a month/4 or more times a week  522 – Prefer not to answer/1 or 2/3 or 4/5 or 6/7,8 or 9/10 or more  523 – Prefer not to anwer/Never/Less than monthly/Monthly/Weekly/Daily or almost daily  524 – Prefer not to answer/Yes, but not in the last year/Yes, during the last year  526 – Prefer not to answer/No/Yes, 1-2 times/Yes, 3-10 times/Yes, 11-100 times/Yes, more than 100 times  532 – Prefer not to answer/Never true/Rarely true/Sometimes true/Often/Very often true  533 – Prefer not to answer/Never/Yes, but not in the last 12 months/Yes, within the last 12 months  534 – Prefer not to answer/Not at all/A little bit/Moderately/Quite a bit/Extremely  535 – Prefer not to answer/No/Yes, once/Yes, more than once  537 – Prefer not to answer/Do not know/Extremely happy/Very happy/Moderately happy/Moderately unhappy/Very unhappy/Extremely unhappy  538 – Prefer not to answer/Do not know/Not at all/A little/A moderate amount/Very much/An extreme amount | | |

| **Phenotype** | **Consortium** | **Sample** | **Citation** | **n_case_** | **n_control_** |
| --- | --- | --- | --- | --- | --- |
| SCZ | PGC | Meta | Pardiñas et al., 2018 | 40,675 | 64,643 |
| BD | PGC | European | Stahl et al., 2019 | 20,352 | 31,358 |
| MDD | PGC | European | Wray et al., 2018 | 69,576 | 161,613 |
| ADHD | PGC | European | Demontis et al., 2019 | 19,099 | 34,194 |
| ASD | PGC, iPSYCH | Meta | Grove et al., 2019 | 18,381 | 27,969 |
| PTSD | PGC | European | Duncan et al., 2018 | 2,424 | 7,113 |
| ANX | ANGST | Meta | Otowa et al., 2016 | 7,016 | 14,745 |
| Intelligence | CTG | Meta | Savage et al., 2018 | 269,867 | |
| Educational attainment | SSGAC | European | Lee et al., 2018 | 766,345 | |
| **Suppl. Table 2.** **Cohort Overview** Overview of the cohorts including references and sample size. | | | | | |

| **IC** | **Component represents** | **h2 (SE)** | **Lambda GC** | **Intercept (SE)** |
| --- | --- | --- | --- | --- |
| IC1 | Sexual abuse | 0.0252 (0.0043) | 1.0557 | 1.0023 (0.0063) |
| IC2 | Psychosis | 0.0089 (0.0043) | 1.0225 | 1.0034 (0.007) |
| IC3 | Anxiety/depression/mental distress | 0.0744 (0.0052) | 1.1523 | 0.9993 (0.0074) |
| IC4 | Diagnosed with life-threatening illness | 0.0181 (0.0041) | 1.0557 | 1.0103 (0.0066) |
| IC5 | Social instability | 0.046 (0.0049) | 1.0988 | 1.0138 (0.0067) |
| IC6 | Traumatic experiences | 0.0313 (0.0041) | 1.0649 | 1.0021 (0.0065) |
| IC7 | Stress last month | 0.0287 (0.0046) | 1.0557 | 0.9978 (0.0068) |
| IC8 | Felt loved | 0.0588 (0.005) | 1.1301 | 1.0058 (0.0067) |
| IC9 | Self-harm | 0.0339 (0.0044) | 1.0741 | 1.0017 (0.0059) |
| IC10 | General happiness | 0.0672 (0.0052) | 1.1555 | 1.0239 (0.0074) |
| IC11 | Addiction/mania | 0.0259 (0.0043) | 1.0710 | 1.0085 (0.0062) |
| IC12 | Emotional abuse | 0.0243 (0.0044) | 1.0527 | 0.996 (0.0062) |
| IC13 | Alcohol abuse | 0.0763 (0.0055) | 1.1587 | 1.0055 (0.0074) |
| **Suppl. Table 3.** **Heritability statistics from LDSC** Heritability estimates and additional statistics on the ICs | | | | |

| **IC** | **Component represents** | **no** | **Gene** | **Chr** | **pmin** | **Individual Significant SNP** |
| --- | --- | --- | --- | --- | --- | --- |
| IC1 | Sexual abuse | 1 | ADARB2 | 10 | 3.74966e-08 | rs554213191 |
| IC1 | Sexual abuse | 2 | ADARB2 | 10 | 3.74966e-08 | rs554213191 |
| IC2 | Psychosis | 1 | GAS2L3 | 12 | 4.49537e-08 | rs540594149 |
| IC2 | Psychosis | 2 | ANO4 | 12 | 7.47504e-07 | rs540594149 |
| IC2 | Psychosis | 3 | FAM109A | 12 | 1.14947e-06 | rs749014627 |
| IC2 | Psychosis | 4 | SH2B3 | 12 | 5.60159e-07 | rs749014627 |
| IC2 | Psychosis | 5 | ATXN2 | 12 | 5.60159e-07 | rs749014627 |
| IC2 | Psychosis | 6 | ACAD10 | 12 | 1.24816e-06 | rs749014627 |
| IC2 | Psychosis | 7 | RP11-162P23.2 | 12 | 1.24816e-06 | rs749014627 |
| IC2 | Psychosis | 8 | ALDH2 | 12 | 1.32971e-06 | rs749014627 |
| IC2 | Psychosis | 9 | MAPKAPK5 | 12 | 5.77870e-07 | rs749014627 |
| IC2 | Psychosis | 10 | TMEM116 | 12 | 1.19305e-05 | rs749014627 |
| IC2 | Psychosis | 11 | HECTD4 | 12 | 4.04123e-06 | rs749014627 |
| IC2 | Psychosis | 12 | GAS2L3 | 12 | 4.49537e-08 | rs540594149 |
| IC2 | Psychosis | 13 | ANO4 | 12 | 7.47504e-07 | rs540594149 |
| IC2 | Psychosis | 14 | FAM109A | 12 | 1.14947e-06 | rs749014627 |
| IC2 | Psychosis | 15 | SH2B3 | 12 | 5.60159e-07 | rs749014627 |
| IC2 | Psychosis | 16 | ATXN2 | 12 | 5.60159e-07 | rs749014627 |
| IC2 | Psychosis | 17 | ACAD10 | 12 | 1.24816e-06 | rs749014627 |
| IC2 | Psychosis | 18 | RP11-162P23.2 | 12 | 1.24816e-06 | rs749014627 |
| IC2 | Psychosis | 19 | ALDH2 | 12 | 1.32971e-06 | rs749014627 |
| IC2 | Psychosis | 20 | MAPKAPK5 | 12 | 5.77870e-07 | rs749014627 |
| IC2 | Psychosis | 21 | TMEM116 | 12 | 1.19305e-05 | rs749014627 |
| IC2 | Psychosis | 22 | HECTD4 | 12 | 4.04123e-06 | rs749014627 |
| IC3 | Anxiety/depression/mental distress | 1 | CABP1 | 12 | 4.21106e-09 | rs73222787 |
| IC3 | Anxiety/depression/mental distress | 2 | MLEC | 12 | 1.62854e-06 | rs73222787 |
| IC3 | Anxiety/depression/mental distress | 3 | UNC119B | 12 | 2.60327e-06 | rs73222787 |
| IC3 | Anxiety/depression/mental distress | 4 | ACADS | 12 | 1.64855e-06 | rs73222787 |
| IC3 | Anxiety/depression/mental distress | 5 | SPPL3 | 12 | 2.82900e-06 | rs73222787 |
| IC3 | Anxiety/depression/mental distress | 6 | CABP1 | 12 | 4.21106e-09 | rs73222787 |
| IC3 | Anxiety/depression/mental distress | 7 | MLEC | 12 | 1.62854e-06 | rs73222787 |
| IC3 | Anxiety/depression/mental distress | 8 | UNC119B | 12 | 2.60327e-06 | rs73222787 |
| IC3 | Anxiety/depression/mental distress | 9 | ACADS | 12 | 1.64855e-06 | rs73222787 |
| IC3 | Anxiety/depression/mental distress | 10 | SPPL3 | 12 | 2.82900e-06 | rs73222787 |
| IC5 | Social instability | 1 | AP1G1 | 16 | 3.00910e-06 | rs8057124 |
| IC5 | Social instability | 2 | ATXN1L | 16 | 1.83621e-08 | rs8057124 |
| IC5 | Social instability | 3 | IST1 | 16 | 1.04493e-08 | rs8057124 |
| IC5 | Social instability | 4 | ZNF821 | 16 | 2.05589e-08 | rs8057124 |
| IC5 | Social instability | 5 | AP1G1 | 16 | 3.00910e-06 | rs8057124 |
| IC5 | Social instability | 6 | ATXN1L | 16 | 1.83621e-08 | rs8057124 |
| IC5 | Social instability | 7 | IST1 | 16 | 1.04493e-08 | rs8057124 |
| IC5 | Social instability | 8 | ZNF821 | 16 | 2.05589e-08 | rs8057124 |
| IC7 | Stress last month | 1 | ACTN1 | 14 | 1.10472e-05 | rs761897 |
| IC7 | Stress last month | 2 | ACTN1 | 14 | 1.10472e-05 | rs761897 |
| IC8 | Felt loved | 1 | EYS | 6 | 4.70962e-08 | rs183356400 |
| IC8 | Felt loved | 2 | RBFOX1 | 16 | 4.39126e-08 | rs13332228 |
| IC8 | Felt loved | 3 | EYS | 6 | 4.70962e-08 | rs183356400 |
| IC8 | Felt loved | 4 | RBFOX1 | 16 | 4.39126e-08 | rs13332228 |
| IC10 | General happiness | 1 | CSMD1 | 8 | 4.77632e-08 | rs2554644 |
| IC10 | General happiness | 2 | CSMD1 | 8 | 4.77632e-08 | rs2554644 |
| IC11 | Addiction/mania | 1 | CADM2 | 3 | 3.68665e-08 | rs9866089 |
| IC11 | Addiction/mania | 2 | CADM2 | 3 | 3.68665e-08 | rs9866089 |
| IC12 | Emotional abuse | 1 | PTPRT | 20 | 1.63897e-08 | rs533568724 |
| IC12 | Emotional abuse | 2 | PTPRT | 20 | 1.63897e-08 | rs533568724 |
| IC13 | Alcohol abuse | 1 | EIF2B4 | 2 | 2.34369e-08 | rs1260326 |
| IC13 | Alcohol abuse | 2 | SNX17 | 2 | 2.34369e-08 | rs1260326 |
| IC13 | Alcohol abuse | 3 | ZNF513 | 2 | 2.34369e-08 | rs1260326 |
| IC13 | Alcohol abuse | 4 | PPM1G | 2 | 2.34369e-08 | rs1260326 |
| IC13 | Alcohol abuse | 5 | GCKR | 2 | 1.70337e-10 | rs1260326 |
| IC13 | Alcohol abuse | 6 | AC109829.1 | 2 | 1.16193e-05 | rs1260326 |
| IC13 | Alcohol abuse | 7 | SIX3 | 2 | 2.32692e-07 | rs528301  rs504675 |
| IC13 | Alcohol abuse | 8 | RFC1 | 4 | 4.44299e-08 | rs12643682 |
| IC13 | Alcohol abuse | 9 | KLB | 4 | 8.52705e-17 | rs12643682  rs58015370  rs13125440  rs6836420 |
| IC13 | Alcohol abuse | 10 | TSPAN5 | 4 | 4.55114e-10 | rs4699663 |
| IC13 | Alcohol abuse | 11 | METAP1 | 4 | 1.26232e-13 | rs146788033 |
| IC13 | Alcohol abuse | 12 | ADH5 | 4 | 3.35425e-25 | rs145452708 |
| IC13 | Alcohol abuse | 13 | ADH6 | 4 | 1.73597e-16 | rs11733695 |
| IC13 | Alcohol abuse | 14 | ADH1B | 4 | 5.89840e-52 | rs1229984  rs145452708 |
| IC13 | Alcohol abuse | 15 | C4orf17 | 4 | 5.76240e-15 | rs543669349 |
| IC13 | Alcohol abuse | 16 | TRMT10A | 4 | 5.76240e-15 | rs543669349 |
| IC13 | Alcohol abuse | 17 | DAPP1 | 4 | NA | rs543669349  rs188514326 |
| IC13 | Alcohol abuse | 18 | LAMTOR3 | 4 | NA | rs543669349  rs188514326 |
| IC13 | Alcohol abuse | 19 | DNAJB14 | 4 | 8.67314e-10 | rs543669349  rs188514326 |
| IC13 | Alcohol abuse | 20 | BANK1 | 4 | 6.49446e-08 | rs13107325 |
| IC13 | Alcohol abuse | 21 | SLC39A8 | 4 | 1.26159e-13 | rs13107325  rs34333163 |
| IC13 | Alcohol abuse | 22 | ARHGAP27 | 17 | 4.06505e-09 | rs62062288 |
| IC13 | Alcohol abuse | 23 | PLEKHM1 | 17 | 1.02001e-13 | rs62062288  rs62053943 |
| IC13 | Alcohol abuse | 24 | CRHR1 | 17 | 1.00879e-13 | rs62062288  rs62053943  rs9303521  rs12944712 |
| IC13 | Alcohol abuse | 25 | SPPL2C | 17 | 1.27409e-13 | rs62062288  rs62053943  rs12944712 |
| IC13 | Alcohol abuse | 26 | MAPT | 17 | 2.31437e-14 | rs62062288  rs62053943 |
| IC13 | Alcohol abuse | 27 | STH | 17 | 1.94674e-13 | rs62062288  rs62053943 |
| IC13 | Alcohol abuse | 28 | KANSL1 | 17 | 5.27802e-14 | rs62062288  rs62053943 |
| IC13 | Alcohol abuse | 29 | ARL17B | 17 | 1.06779e-13 | rs62062288  rs62053943 |
| IC13 | Alcohol abuse | 30 | LRRC37A | 17 | 1.06779e-13 | rs62062288  rs62053943 |
| IC13 | Alcohol abuse | 31 | NSF | 17 | 9.06747e-13 | rs62062288  rs62053943 |
| IC13 | Alcohol abuse | 32 | WNT3 | 17 | 7.33108e-12 | rs62062288 |
| IC13 | Alcohol abuse | 33 | FUT2 | 19 | 2.24286e-08 | rs28894750 |
| IC13 | Alcohol abuse | 34 | MAMSTR | 19 | 2.24286e-08 | rs28894750 |
| IC13 | Alcohol abuse | 35 | RASIP1 | 19 | 4.60338e-08 | rs28894750 |
| IC13 | Alcohol abuse | 36 | IZUMO1 | 19 | 2.76726e-06 | rs28894750 |
| IC13 | Alcohol abuse | 37 | FUT1 | 19 | 2.76726e-06 | rs28894750 |
| IC13 | Alcohol abuse | 38 | EIF2B4 | 2 | 2.34369e-08 | rs1260326 |
| IC13 | Alcohol abuse | 39 | SNX17 | 2 | 2.34369e-08 | rs1260326 |
| IC13 | Alcohol abuse | 40 | ZNF513 | 2 | 2.34369e-08 | rs1260326 |
| IC13 | Alcohol abuse | 41 | PPM1G | 2 | 2.34369e-08 | rs1260326 |
| IC13 | Alcohol abuse | 42 | GCKR | 2 | 1.70337e-10 | rs1260326 |
| IC13 | Alcohol abuse | 43 | AC109829.1 | 2 | 1.16193e-05 | rs1260326 |
| IC13 | Alcohol abuse | 44 | SIX3 | 2 | 2.32692e-07 | rs528301  rs504675 |
| IC13 | Alcohol abuse | 45 | RFC1 | 4 | 4.44299e-08 | rs12643682 |
| IC13 | Alcohol abuse | 46 | KLB | 4 | 8.52705e-17 | rs12643682  rs58015370  rs13125440  rs6836420 |
| IC13 | Alcohol abuse | 47 | TSPAN5 | 4 | 4.55114e-10 | rs4699663 |
| IC13 | Alcohol abuse | 48 | METAP1 | 4 | 1.26232e-13 | rs146788033 |
| IC13 | Alcohol abuse | 49 | ADH5 | 4 | 3.35425e-25 | rs145452708 |
| IC13 | Alcohol abuse | 50 | ADH6 | 4 | 1.73597e-16 | rs11733695 |
| IC13 | Alcohol abuse | 51 | ADH1B | 4 | 5.89840e-52 | rs1229984  rs145452708 |
| IC13 | Alcohol abuse | 52 | C4orf17 | 4 | 5.76240e-15 | rs543669349 |
| IC13 | Alcohol abuse | 53 | TRMT10A | 4 | 5.76240e-15 | rs543669349 |
| IC13 | Alcohol abuse | 54 | DAPP1 | 4 | NA | rs543669349  rs188514326 |
| IC13 | Alcohol abuse | 55 | LAMTOR3 | 4 | NA | rs543669349  rs188514326 |
| IC13 | Alcohol abuse | 56 | DNAJB14 | 4 | 8.67314e-10 | rs543669349  rs188514326 |
| IC13 | Alcohol abuse | 57 | BANK1 | 4 | 6.49446e-08 | rs13107325 |
| IC13 | Alcohol abuse | 58 | SLC39A8 | 4 | 1.26159e-13 | rs13107325  rs34333163 |
| IC13 | Alcohol abuse | 59 | ARHGAP27 | 17 | 4.06505e-09 | rs62062288 |
| IC13 | Alcohol abuse | 60 | PLEKHM1 | 17 | 1.02001e-13 | rs62062288  rs62053943 |
| IC13 | Alcohol abuse | 61 | CRHR1 | 17 | 1.00879e-13 | rs62062288  rs62053943  rs9303521  rs12944712 |
| IC13 | Alcohol abuse | 62 | SPPL2C | 17 | 1.27409e-13 | rs62062288  rs62053943  rs12944712 |
| IC13 | Alcohol abuse | 63 | MAPT | 17 | 2.31437e-14 | rs62062288  rs62053943 |
| IC13 | Alcohol abuse | 64 | STH | 17 | 1.94674e-13 | rs62062288  rs62053943 |
| IC13 | Alcohol abuse | 65 | KANSL1 | 17 | 5.27802e-14 | rs62062288  rs62053943 |
| IC13 | Alcohol abuse | 66 | ARL17B | 17 | 1.06779e-13 | rs62062288  rs62053943 |
| IC13 | Alcohol abuse | 67 | LRRC37A | 17 | 1.06779e-13 | rs62062288  rs62053943 |
| IC13 | Alcohol abuse | 68 | NSF | 17 | 9.06747e-13 | rs62062288  rs62053943 |
| IC13 | Alcohol abuse | 69 | WNT3 | 17 | 7.33108e-12 | rs62062288 |
| IC13 | Alcohol abuse | 70 | FUT2 | 19 | 2.24286e-08 | rs28894750 |
| IC13 | Alcohol abuse | 71 | MAMSTR | 19 | 2.24286e-08 | rs28894750 |
| IC13 | Alcohol abuse | 72 | RASIP1 | 19 | 4.60338e-08 | rs28894750 |
| IC13 | Alcohol abuse | 73 | IZUMO1 | 19 | 2.76726e-06 | rs28894750 |
| IC13 | Alcohol abuse | 74 | FUT1 | 19 | 2.76726e-06 | rs28894750 |
| **Suppl. Table 4.** **Mapped genes from FUMA** This table shows the mapped genes and individual lead SNPs as provided by FUMA. | | | | | | |

**Supplementary Figures**

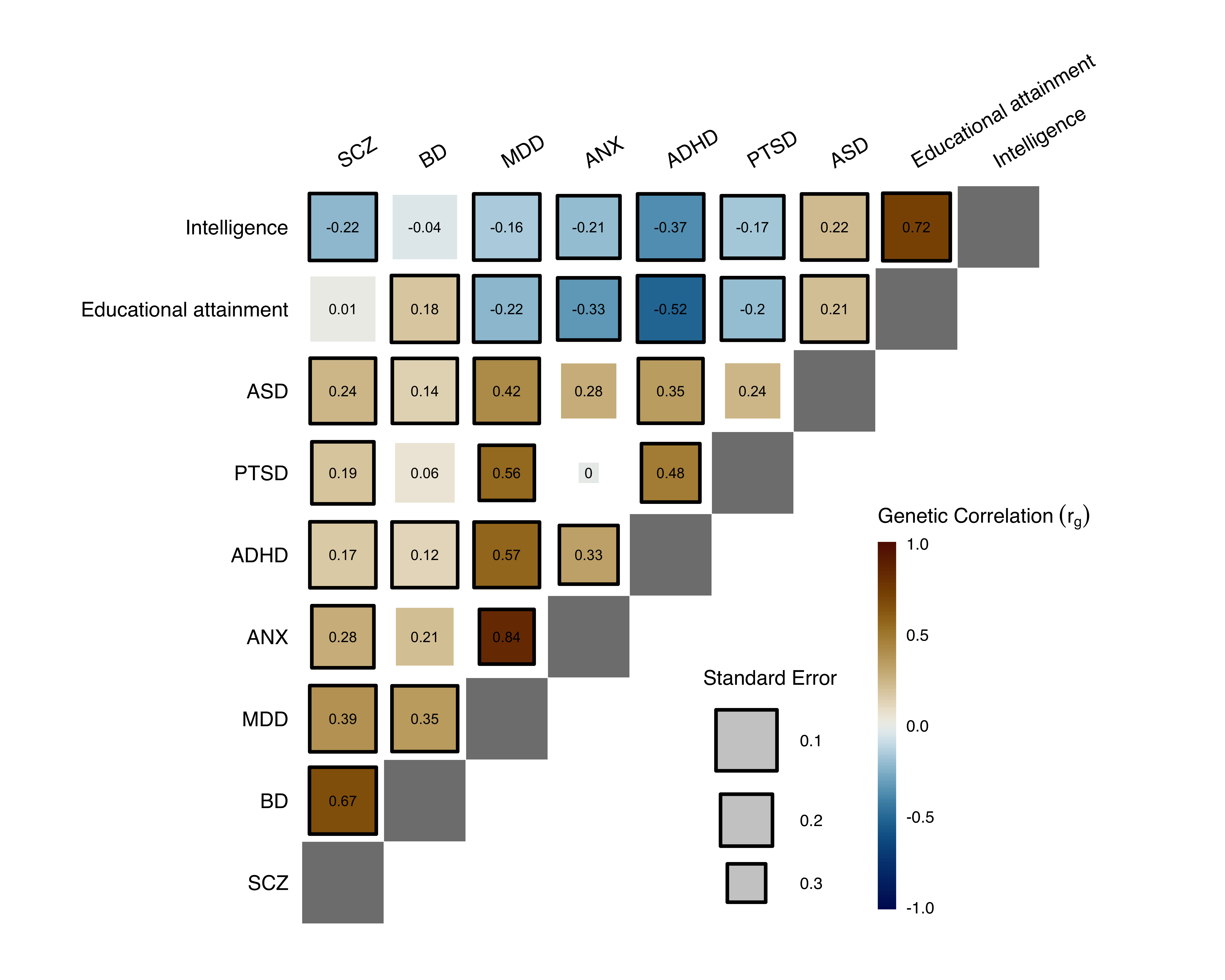

**Suppl. Fig. 1.** **Genetic correlation between the disorders and cognitive traits** Numbers inside the boxes denote correlation (r_g_). Size of the boxes reflect standard error. Significant correlations (p < FDR) are indicated with a black border. In line with previous reports^9,30^, the weakest correlation was between PTSD and ANX (r_g_ = -0.004, SE = 0.3408) and the strongest between ANX and MDD (r_g_ = 0.8441, SE = 0.1724).

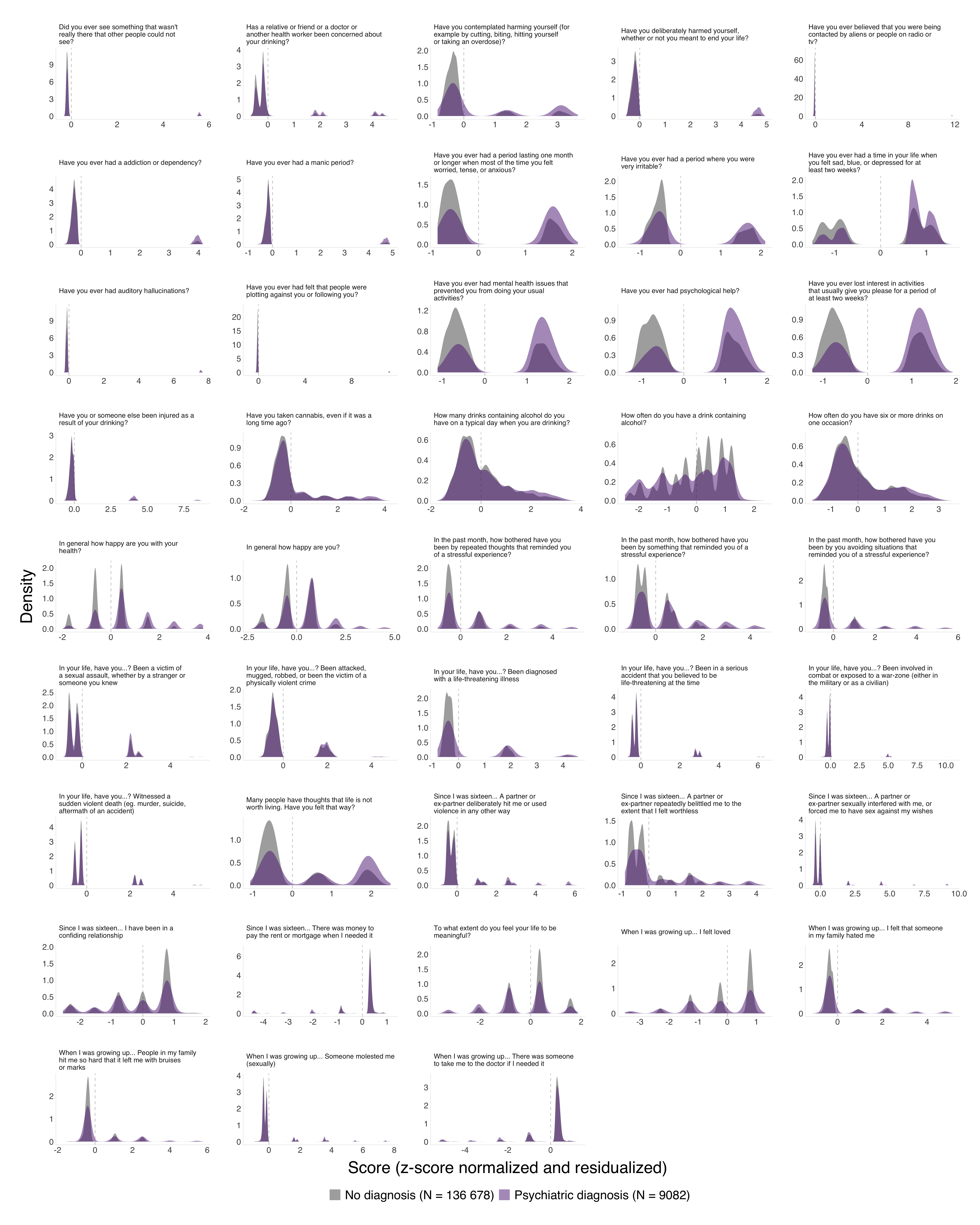

**Suppl. Fig. 2. Z-normalized question scores** Density plot of the scores comparing the sample of individuals without psychiatric or neurological diagnosis to the individuals with a diagnosed psychiatric disorder. With a few notable exceptions, distributions are fairly similar, supporting that mental health data from healthy individuals can be used to study psychiatric disorders. As expected, in the exceptions where distributions were different, individuals with a psychiatric diagnosis had higher scores overall compared to healthy individuals.

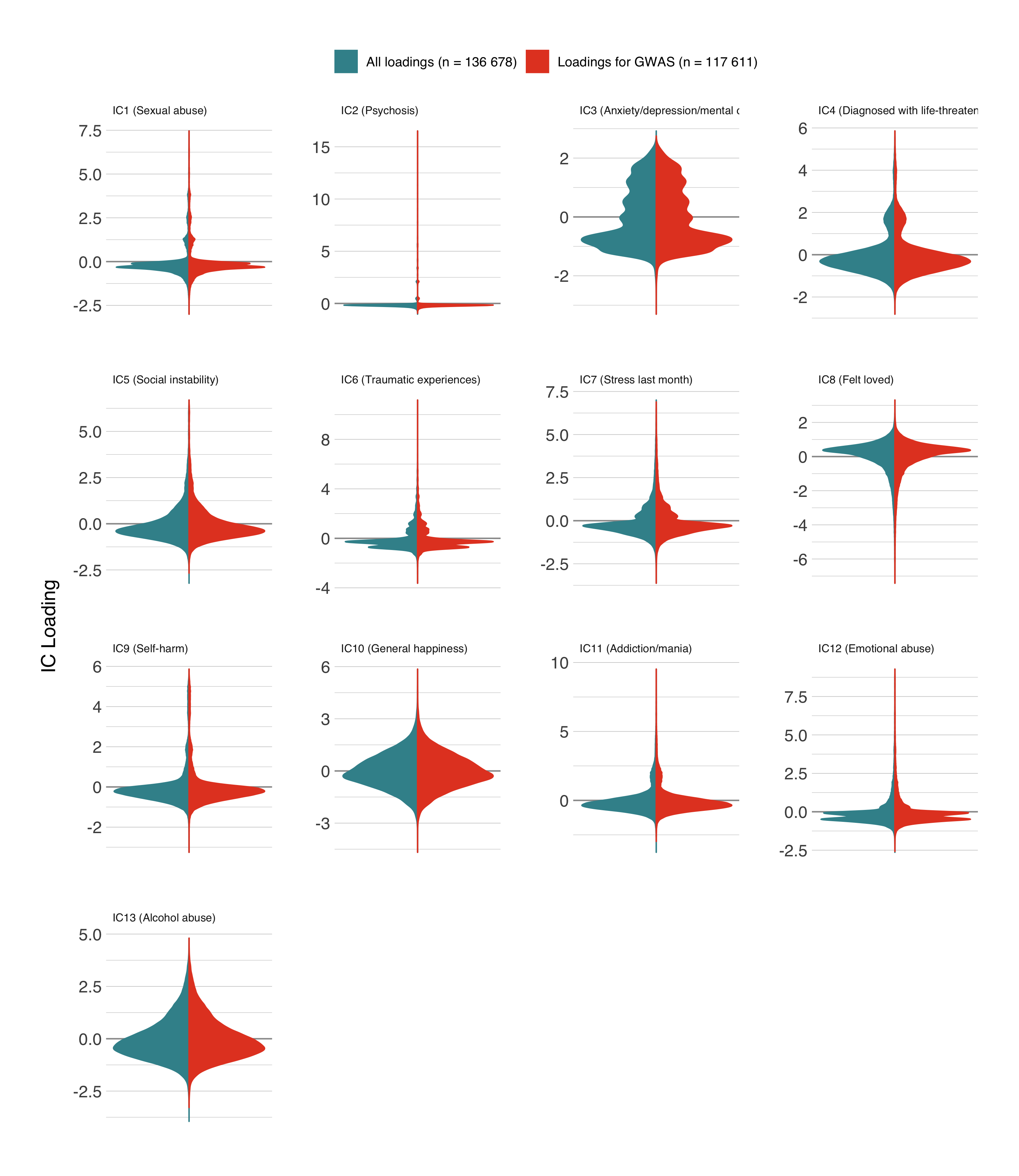

**Suppl. Fig. 3.** **ICA weight distributions** Density plot of the individual loadings for each independent component. Left half of the plot indicate the loadings for all individuals that were used for ICA. The right side reflects the sample from which genetic data was available and these are the loadings that were used for the GWASs. Distributions on the left show the loadings as provided by the ICA. Distributions were regressed for age (linear and quadratic), sex, and the first 20 genetic principal components before ICA decomposition. Loadings of IC1, IC2, IC5, IC9, IC10, IC11, and IC12 were inverted so that higher loadings reflect higher symptoms. IC2 has a very narrow distribution, with a few high loadings. IC10 showed a wide distribution due to the universal nature of the questions that comprise this component.

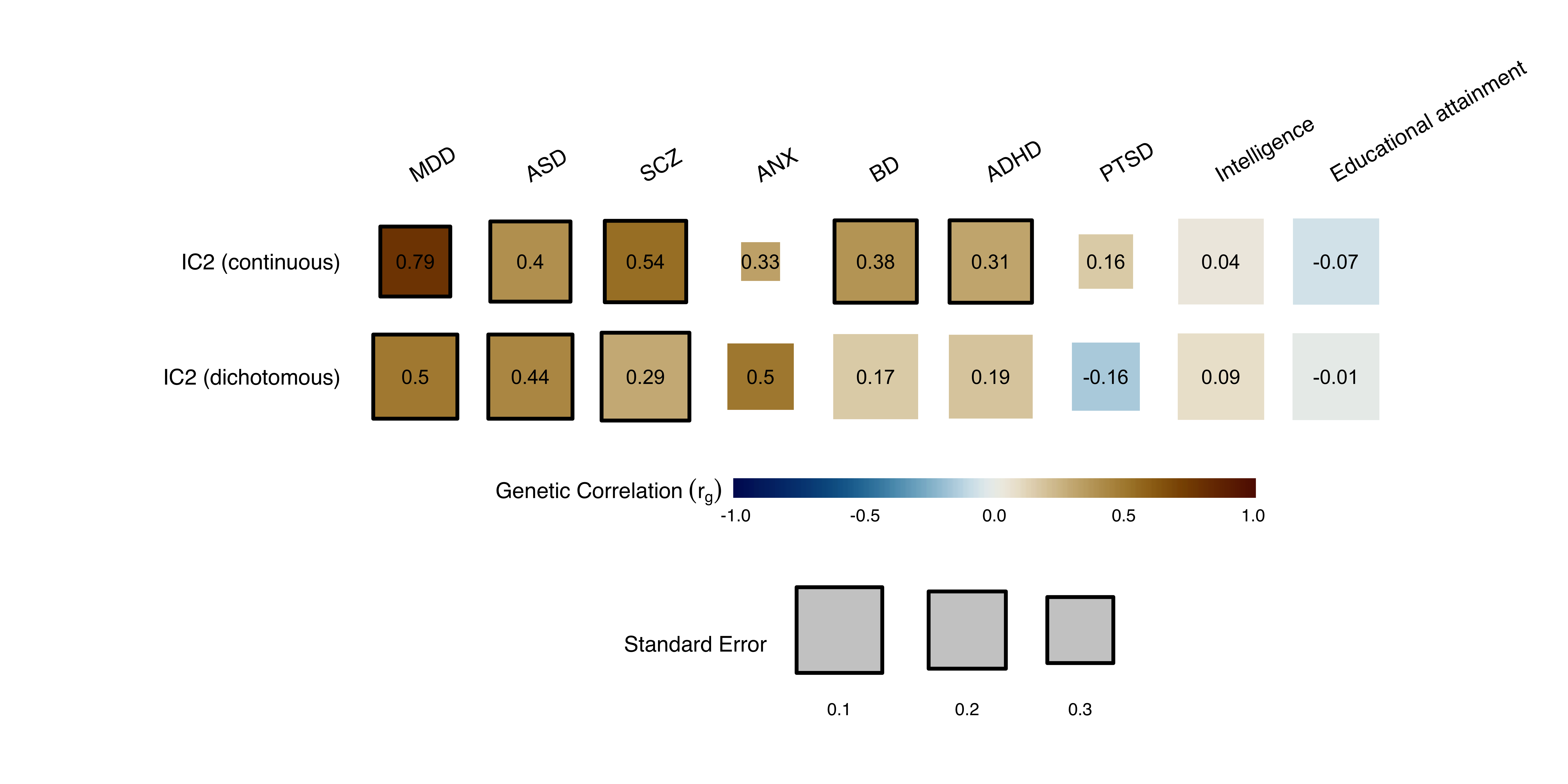

**Suppl. Fig. 4.** **Comparison between continuous and dichotomous IC2** Due to the low variance in IC2 particularly (Suppl. Fig. 3), we investigated if dichotomizing the loading on that component would improve heritability (i.e. loadings larger than 1 were used as cases, lower than 1 were used as controls in a case-control GWAS). This dichotomous approach is conceptually similar to the approach taken by Legge and colleagues^30^ who recently reported a genetic correlation between SCZ and dichotomous psychosis symptoms (yes/no). However, dichotomizing did not improve the estimates strongly. Heritability was only slightly higher (h2_dichotomous_ = 0.0133 vs h2_continuous_ = 0.0089) and the reported association with SCZ decreased from r_g continuous_ = 0.5427 to r_g_ _dichotomous_ = 0.2945. The standard error was more than halved after dichotomization (SE_continuous_ = 0.1527 vs SE_dichotomous_ = 0.0711), suggesting a slight benefit of the dichotomization in scenarios where data distributions limit the continuous approach. In the main analyses, we decided to stay consistent with the other components and keep the phenotype as a continuous measure for IC2, yet we provide results with the dichotomized component for comparison in this figure. Correlations are mostly the same for the continuous and the dichotomous component. The continuous IC2 correlated stronger with MDD compared to the dichotomous IC2, which correlated less strongly with MDD, but stronger with ASD and ANX compared to the continuous IC2. Errors were relatively similar.

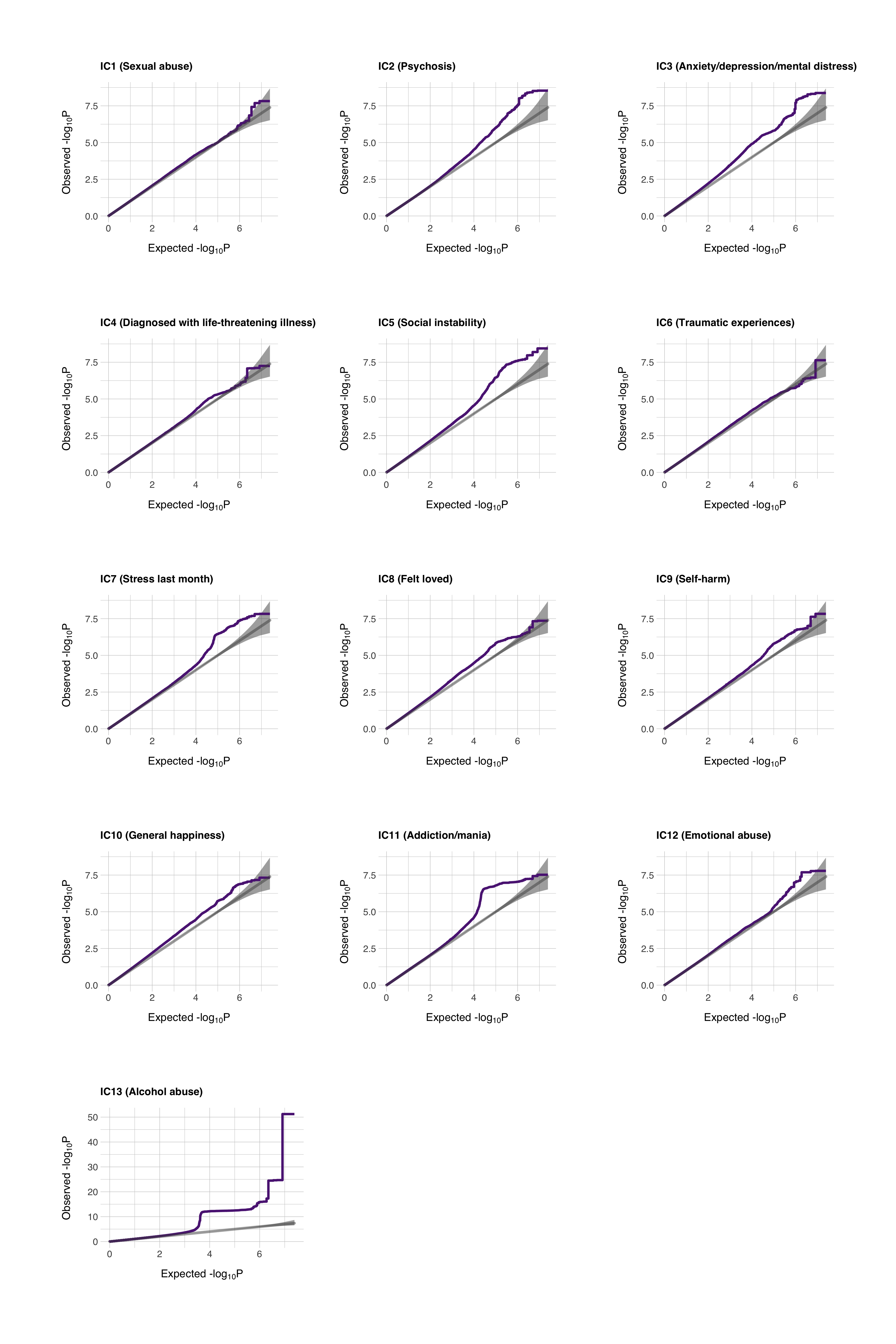

**Suppl. Fig. 5.** **Q-Q plots** Q-Q plots showing the genetic signal from each of the ICs. IC13 showed the strongest signal. IC2 showed a strong signal despite having the lowest h2. None of the GWAS summary statistics showed any noticeable inflation.

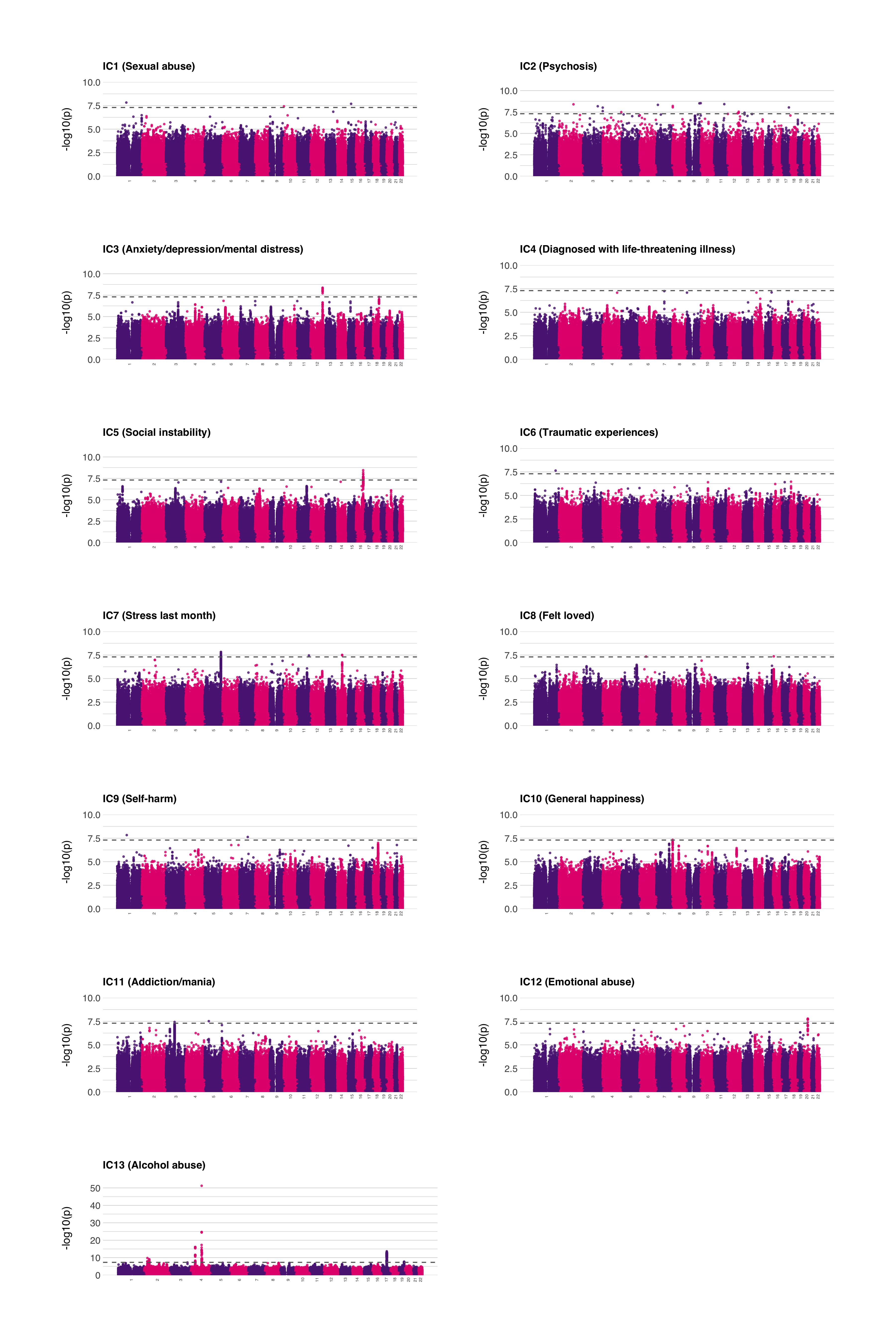

**Suppl. Fig. 6.** **Manhattan plots for the ICs** Significance threshold set at 5e-8. Only IC4 had no genome-wide significant hits, a few more had no lead SNPs, according to FUMA.

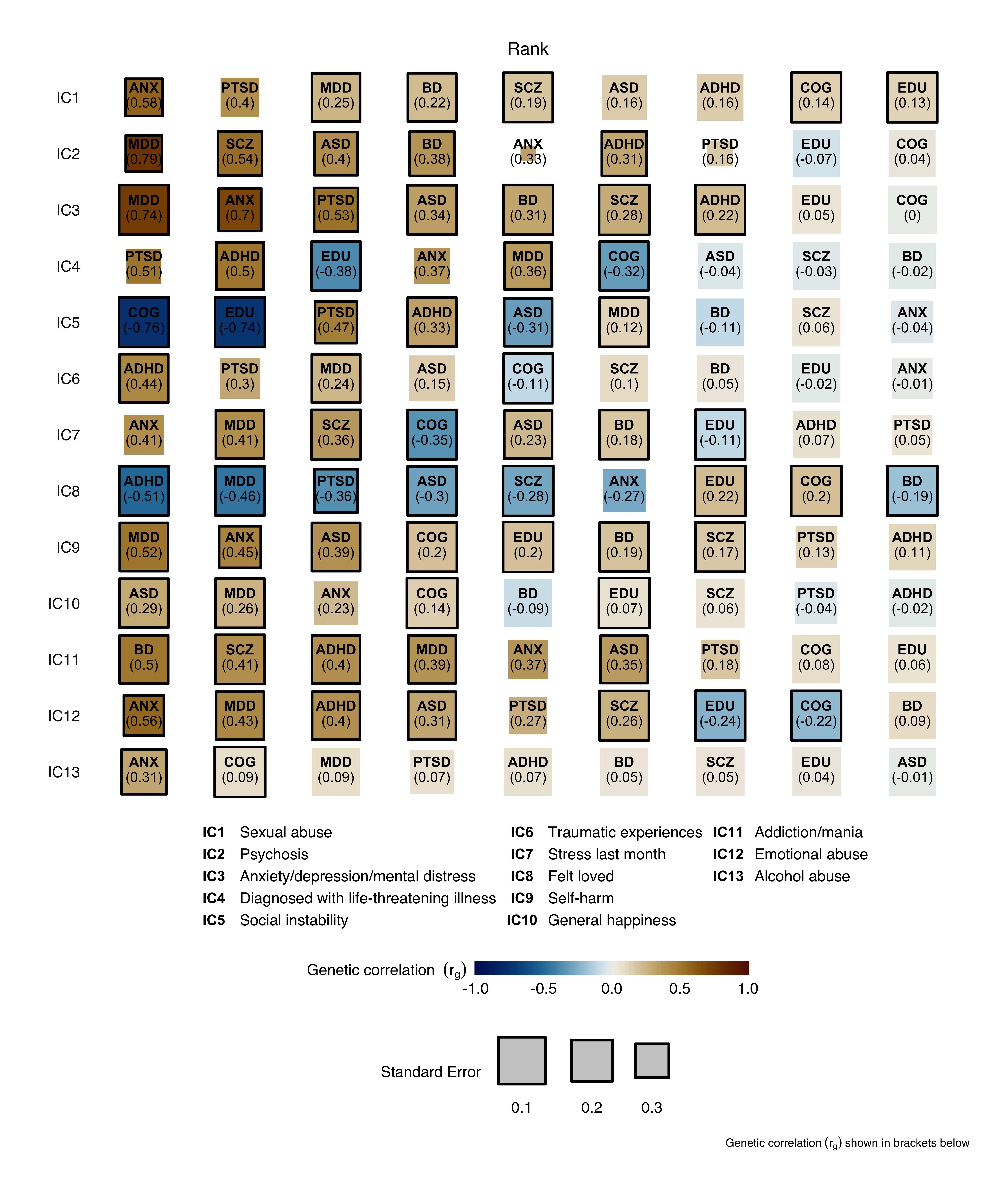

**Suppl. Fig. 7.** **Genetic correlation between the independent components and disorders and cognitive traits separated by independent component** As opposed to of Fig. 3 where correlations are separated by psychiatric diagnosis or cognitive trait, this figure shows the genetic correlations between the independent components and the psychiatric diagnoses and cognitive traits separated by independent component. The horizontal ordering of the tiles corresponds to the strength of the correlation (r_g_) so that the left-most tile is the strongest correlation (either negative or positive) and the right-most tile shows the lowest correlation.
